# Topological Aberrance of Structural Brain Network Provides Quantitative Markers of post-TBI Attention Deficits in Children

**DOI:** 10.1101/2020.06.12.20129890

**Authors:** Meng Cao, Yuyang Luo, Ziyan Wu, Catherine A. Mazzola, Arlene Goodman, Lori Catania, Tara L. Alvarez, Jeffrey M. Halperin, Xiaobo Li

## Abstract

Traumatic brain injury (TBI)-induced attention deficits are among the most common long-term cognitive consequences in children. Most of the existing studies attenpting to understand the neuropathological underpinnings of cognitive and behavioral impairments in TBI have utilized heterogeneous samples and resulted in inconsistent findings. The current research proposed to investigate topological properties of the structural brain network in children with TBI and their associations with TBI-induced attention problems in a more homogeneous subgroup of children who had severe post-TBI attention deficits (TBI-A).

A total of 31 children with TBI-A and 35 group-matched controls were involved in the study. Diffusion tensor imaging-based probabilistic tractography and graph theoretical techniques were used to construct the structural brain network in each subject. Network topological properties were calculated in both global level and regional (nodal) level. Between-group comparisons among the topological network measures and analyses for searching brain-behavioral associations were all corrected for multiple comparisons using Bonferroni method.

Compare to controls, the TBI-A group showed significantly higher nodal local efficiency and nodal clustering coefficient in left inferior frontal gyrus and right transverse temporal gyrus, while significantly lower nodal clustering coefficient in left supramarginal gyrus as well as lower nodal local efficiency in left parahippocampal gyrus. The temporal lobe topological alterations were significantly associated with the post-TBI inattentive and hyperactive symptoms in the TBI-A group.

The results suggest that TBI-related structural re-modularity in the WM subnetworks associated with temporal lobe may play a critical role in the onset of severe post-TBI attention deficits in children. These findings provide valuable input for understanding the neurobiological substrates of TBI-A, and have the potential to serve as a biomarker guiding the development of more timely and tailored strategies for diagnoses and treatments to the affected individuals.

## 1. Introduction

Pediatric traumatic brain injury (TBI) is a major public health concern, which occurs in more than 100,000 children each year and incurs an estimated annual medical cost of more than $1 billion (Watson et al., 2019). Neurocognitive impairments and behavioral abnormalities have been consistently observed in children with TBI (Konigs et al., 2015;Poslinder et al., 2015;Dewan et al., 2016;Lumba-Brown et al., 2018). Among the most common cognitive consequences, significant attention deficits were reported in about 35% of children within two years of their TBI (Max et al., 2005), and were observed to strongly contribute to elevated risk for severe psychopathology and impairments in overall functioning in late adolescence, with the pathophysiological underpinning yet to be fully elucidated (Le Fur et al., 2019;Narad et al., 2019). The post-TBI attention problems in children have been evaluated and treated based on endorsements of behavioral symptoms from subjective observations, and have resulted in largely divergent results regarding effectiveness (Backeljauw and Kurowski, 2014;Kurowski et al., 2019;LeBlond et al., 2019). Understanding the neurobiological substrates of post-TBI attention deficits in children is thus vitally critical, so that timely and tailored strategies can be developed for diagnoses and long-term treatments and interventions.

In literature of pediatric TBI, injury-induced regional structural brain alterations and associated cognitive and behavioral impairments have been increasingly reported. Structural MRI studies have investigated the relationship between cortical thickness and functional outcomes in children with chronic TBI, and found that the abnormal cortical gray matter (GM) thickness in frontal, parietal, and temporal regions were significantly associated with working memory impairments (Merkley et al., 2008) and executive dysfunctions (Wilde et al., 2012b). Existing diffusion tensor imaging (DTI) studies in children with TBI have also reported widespread white matter (WM) structural abnormalities and their associations with post-TBI cognitive and behavioral impairments in the chronic stage. For instance, a number of DTI studies have demonstrated that disrupted WM integrity in corpus collosum (Ewing-Cobbs et al., 2008;Wilde et al., 2011;Treble et al., 2013;Lindsey et al., 2019), uncinate fasciculus (Lindsey et al., 2019), superior longitudinal fasciculus and inferior fronto-occipital fasciculus (Dennis et al., 2015) were significantly associated with working memory impairments in children with TBI. Lower fractional anisotropy (FA) in frontal regions (Wozniak et al., 2007;Kurowski et al., 2009), superior longitudinal fasciculus and anterior corona radiata (Adamson et al., 2013), and ventral striatum (Faber et al., 2016) have been found to significantly link to post-TBI executive dysfunctions in children. Reduced FA in inferior longitudinal fasciculus, inferior fronto-occipital fasciculus, superior longitudinal fasciculus and corpus collosum were also found to be associated with impaired attention function (Konigs et al., 2018). The large inconsistency of these findings was partially resulted from factors of the study samples, such as heterogeneity regarding TBI-induced cognitive and behavioral impairments and their severity levels, variations in terms of the biological and modifiable factors, differences in injury severity and mechanism, sample sizes, differences in imaging and data analysis techniques, etc. In addition, for understanding relations of the anatomical and cognitive/behavioral alterations in TBI, the region-of-interest (ROI)-based investigations of the injured human brain can be biased without considering the fact that human brain is formed as a structurally and functionally connected network for information-transferring.

Indeed, human brain regions do not work in an isolated manner. When processing sensory and higher-order cognitive information, cortical and subcortical brain regions have been found to dynamically reassemble into small-world networks, to maintain optimal communication efficiency (Bassett et al., 2011;Spreng et al., 2013). Structural connectome, facilitated by WM structural connectivity, has been highlighted to play important role in supporting functional brain processes (Baum et al., 2017;Chu et al., 2018). In the context of TBI in children, diffuse injury of the WM has been demonstrated to damage the structural networks, resulting in disrupted functional information transfer across distal brain regions and cognitive and behavioral impairments. For instance, structural brain network studies have found that, relative to matched controls, children with chronic TBI had altered global network properties, including increased characteristic path length and decreased local efficiency, suggesting a more segregated, instead of a normally more coordinated, architecture for information processing (Caeyenberghs et al., 2012;Konigs et al., 2017;Yuan et al., 2017). In addition, the reduced connectivity in the network was found to associate with deficits in postural control (Caeyenberghs et al., 2012), and decreased IQ and impaired working memory (Konigs et al., 2017) in TBI children. A longitudinal intervention study reported that improved overall cognitive performance after intervention was associated reduced network segregation in TBI children (Yuan et al., 2017). A more recent study categorized the edges of the structural brain network into rich club (connections between different hubs), feeder (connections between hubs and other nodes), and local (connections between different non-hub nodes) connections, and reported that children with TBI had significantly lower overall strength in rich club connections and higher overall strength in local connections; while none were associated with their significantly impaired executive function (Verhelst et al., 2018). Although increasing number of studies have started to focus their effort on understanding the relations of TBI-induced structural brain network alterations and cognitive/behavioral impairments (Imms et al., 2019), the neuroanatomical substrates of severe post-TBI attention deficits in children have not yet been fully investigated.

The current study proposed to utilize the probabilistic tractography in DTI and graph theoretical techniques to assess the structural connectome properties in a carefully evaluated cohort of children with severe post-TBI attention deficits and group-matched controls. In previous functional MRI studies, significant functional hyper-activations in frontal and parietal regions have been consistently observed in children with TBI, during sustained attention and inhibitory control processes (Kramer et al., 2008;Tlustos et al., 2011;Strazzer et al., 2015). Based on these findings, we hypothesized that altered regional structural network properties in frontal and parietal areas may exist in children with severe post-TBI attention deficits.

## 2. Materials and methods

### 2.1. Participants

A total of 66 children, including 31 with severe post-TBI attention deficits (TBI-A) and 35 group-matched controls, were initially involved in this study. A subject in the TBI-A group must have had a clinically diagnosed mild or moderate non-penetrating TBI at least 6 months prior to the study date; and T score ≥ 65 in the inattention subscale (or T scores ≥ 65 in both inattention and hyperactivity subscales) in the Conners 3^rd^ Edition-Parent Short form (Conners 3-PS) (Conners, 2008) assessed during the study visit. Children with TBI who had overt focal brain damages or hemorrhages were exclusive.

To rule out confounding factors associated with pre-TBI attention deficits, children who had a history of diagnosed ADHD (any sub-presentations) prior the diagnosis of TBI, or severe pre-TBI inattentive and/or hyperactive behaviors that were reported by a parent, were exclusive from the TBI-A group. The control group included children with no history of diagnosed TBI, no history of diagnosed ADHD, and T scores ≤ 60 in all of the subscales in the Conners 3-PS assessed during the study visit.

To further improve the homogeneity of the study sample, the general inclusion criteria for both groups included 1) only right-handed, to remove handedness-related potential effects on brain structures; 2) full scale IQ ≥ 80, to minimize neurobiological heterogeneities in the study sample; 3) ages of 11 – 15 years, to reduce neurodevelopment-introduced variations in brain structures. In the study, handedness was evaluated using the Edinburgh Handedness Inventory (Oldfield, 1971). Full scale IQ was estimated by the Wechsler Abbreviated Scale of Intelligence II (WASI-II) (Wechsler, 2011). The two groups were matched on sex (male/female) distribution and socioeconomic status (SES) that was estimated using the average education year of both parents.

The general exclusion criteria for both groups were 1) current or previous diagnosis of Autism Spectrum Disorders, Pervasive Development Disorder, psychotic, Major Mood Disorders (except dysthymia not under treatment), Post-Traumatic Stress Disorder, Obsessive-Compulsive Disorder, Conduct Disorder, Anxiety (except simple phobias), or substance use disorders, based on Diagnostic and Statistical Manual of Mental Disorders 5 (DSM-5) (Association, 2013) and supplemented by the Kiddie Schedule for Affective Disorders and Schizophrenia for School-Age Children-Present and Lifetime Version (K-SADS-PL) (Kaufman et al., 2000); 2) any types of diagnosed chronic medical illnesses, neurological disorders, or learning disabilities, from the medical history; 3) treatment with long-acting stimulants or non-stimulant psychotropic medications within the past month; 4) any contraindications for MRI scanning, such as claustrophobia, tooth braces or other metal implants; 5) pre-puberty subjects were also exclusive, to reduce confounders associated with different pubertal stages (Blakemore and Choudhury, 2006). Puberty status was evaluated using the parent version of Carskadon and Acebo’s self-administered rating scale (Carskadon and Acebo, 1993).

After initial processing of the neuroimaging data from each subject, 3 subjects were excluded from further analyses, due to heavy head motion. Therefore, a total of 31 patients with TBI-A and 32 controls were included in group-level analyses.

The TBI-A subjects were recruited from the New Jersey Pediatric Neuroscience Institute (NJPNI), North Jersey Neurodevelopmental Center (NJNC), Children’s Specialized Hospital (CSH), Brain Injury Alliance of New Jersey (BIANJ), and local communities in New Jersey. Controls were solicited from the local communities by advertisement in public places. The study received institutional review board approval at the New Jersey Institute of Technology (NJIT), Rutgers University, and Saint Peter’s University Hospital. Prior the study, all the participants and their parents or guardians provided written informed assents and consents, respectively.

### 2.2. Clinical/Neurocognitive Assessments and Measures

Severity of TBI was characterized using the Glasgow Coma Scale (GCS) (Teasdale and Jennett, 1974), with the scores ranging from 9 to 15 in the TBI-A subjects. Severities of the inattentive and hyperactive/impulsive symptoms were dimensionally measured using the raw scores and T scores of the subscales in Conners 3-PS. The CogState brief battery for children (Eckner et al., 2011), which included 5 computerized tests, was administered to each subject. The normalized overall scores of the tests were used to evaluate neurocognitive capacities in executive function, psychomotor speed, visual attention, visual learning/memory and working memory.

All of the demographic, clinical, and neurocognitive performance measures were summarized in **Table 1**.

**Table 1.** Demographic, clinical, and neurocognitive characteristics in the study sample. TBI-A: TBI induced attention deficit; SD: Standard deviation; N: Number of subjects; M: Males; F: Females.

|  | **Controls**<br>**Mean (SD)** | **TBI-A**<br>**Mean (SD)** | **t or $\chi^2$ value** | **p value** |
| --- | --- | --- | --- | --- |
| **N** | 35 (M:18, F:17) | 31 (M:16, F:15) | 0.0002 ($\chi^2$) | 0.988 |
| **Age** | 13.83 (1.29) | 14.13 (1.69) | -0.817 | 0.417 |
| **Ethnicity/Race** | | | 0.2418 ($\chi^2$) | 0.886 |
| Caucasian | 25 | 23 |  |  |
| Hispanic | 6 | 4 |  |  |
| Others | 4 | 4 |  |  |
| **Socio-Economic Status** | 32.00 (3.70) | 31.16 (4.16) | 0.872 | 0.387 |
| **Full Scale IQ** | 115.08 (11.27) | 111.13 (12.59) | 1.348 | 0.183 |
| **Conners 3rd Edition-Parent Short Form (T-score)** |  |  |  |  |
| Inattention | 45.97 (5.86) | 70.52 (7.32) | -18.031 | < 0.001 |
| Hyperactivity/Impulsivity | 48.31 (5.55) | 62.00 (14.08) | -6.546 | < 0.001 |
| **CogState Brief Neurocognitive Testing Battery** |  |  |  |  |
| Groton Maze Learning Test | 106.34 (4.76) | 105.58 (4.71) | 0.653 | 0.516 |
| Detection | 100.54 (6.02) | 101.39 (4.96) | -0.617 | 0.540 |
| Identification | 102.17 (5.25) | 100.39 (6.68) | 1.213 | 0.230 |
| One Card Learning | 101.34 (7.36) | 100.16 (7.53) | 0.644 | 0.522 |
| One Back (Speed) | 92.63 (9.18) | 93.58 (10.61) | -0.391 | 0.697 |
| One Back (Accuracy) | 101.77 (6.94) | 104.55 (8.20) | -1.490 | 0.141 |

### 2.3. Neuroimaging Data Acquisition Protocol

MRI scans for each subject were performed on a 3-Tesla Siemens TRIO (Siemens Medical Systems, Germany) scanner at Rutgers University Brain Imaging Center. The DTI data were acquired using a single-shot echo planar sequence (voxel size=2.0 mm × 2.0 mm × 2.5 mm voxel size, repetition time (TR)=7700 ms, echo time (TE) = 103 ms, field of view (FOV) = 250 mm × 250 mm, 30 diffusion-sensitizing gradient directions with b-value = 700 s/mm^2^, and one image with b-value = 0 s/mm^2^). In addition, high-resolution T1-weighted data from each subject was also involved in the study for creation of individualized brain atlas. The T1-weighted anatomical images were obtained with a sagittal multi-echo magnetization-prepared rapid acquisition gradient echo (MPRAGE) sequence (voxel size=1 mm^3^ isotropic, TR = 1900 ms, TE = 2.52 ms, flip angle = 9°, FOV = 250 mm × 250 mm, and 176 sagittal slices).

### 2.4. Individual-level Neuroimaging Data Pre-processes

DTI data pre-processing was performed using the Diffusion Toolbox from FMRIB Software Library v6.0 (FSL) (Jenkinson et al., 2012). Each DTI data (**Figure 1A** as an example) was first manually checked for any missing slides or heavy geometric distortions. The head-motions and eddy-current distortion were then corrected with affine transformation and predictions estimated by a Gaussian Process (Andersson and Sotiropoulos, 2016). The cutoff of heavy head movements was defined as 2mm translational motion in any of the three directions, with which data from 3 subjects were excluded from further analyses. Non-brain voxels were removed by performing brain extraction over the non-diffusion-weighted image (b0 image). The parameters for probabilistic tractography were estimated using the FSL/BedpostX toolbox (Behrens et al., 2007). This process estimated a 2-fiber model in each voxel based on the probability distribution generated by Markov Chain Monte Carlo sampling (**Figure 1B**).

**Figure 1.**
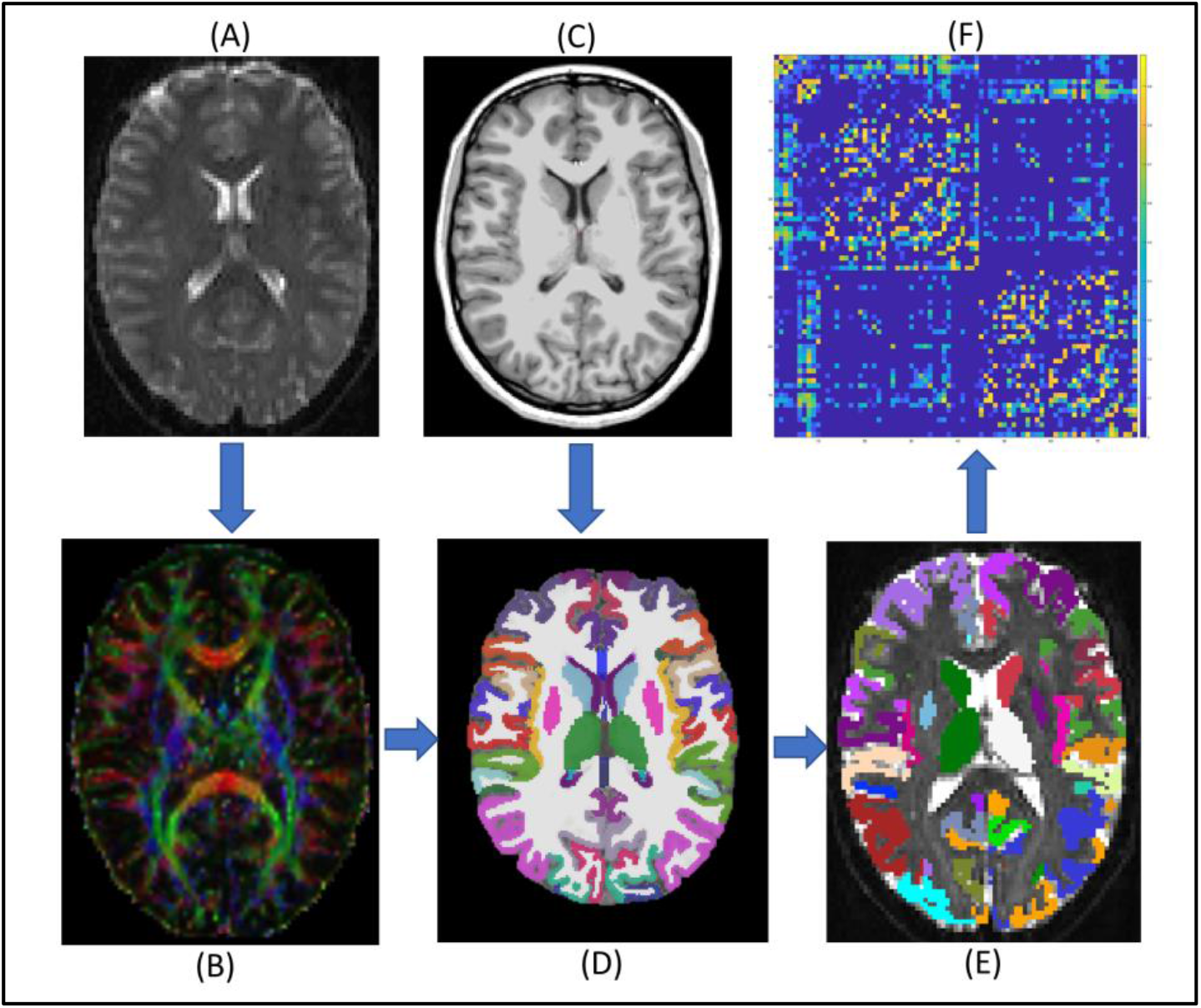
Individual level imaging data analysis and network construction. (**A**) DTI data; (**B**) Estimated tensor directions with 2-crossing fiber model; (**C**) T1-weighted structural MRI data; (**D**) Parcellated structural image based on Desikan-Killiany Atlas; (**E**) Seed masks in diffusion space, which binarized and transformed from structural space; (F) Symmetric and weighted 78 × 78 connectivity matrix. The edges were calculated based on the number of fibers in tractography. DTI: Diffusion Tensor Imaging.

In each subject, a total of 78 cortical and subcortical ROIs were generated from the T1-weighted data (**Figure 1C**) using the standardized brain atlas parcellation procedures from FreeSurfer v6.0.0 (Fischl, 2012). There ROIs (**Figure 1D**) included 68 cortical regions bilaterally, and 10 subcortical regions (bilateral putamen, caudate, hippocampus, thalamus, and pallidum). All of the ROIs in structural space were linearly registered into each individual’s native diffusion space by referencing to the b0 image and binarized into ROI masks to serve as seed masks for tractography (**Figure 1E**).

Finally, the DTI probabilistic fiber tracking was performed using a streamline tractography algorithm, FSL/PROBTRACKX2. To prevent the generated fibers from running into GM and cerebrospinal fluid, a WM mask was used for the probabilistic tractography. Five thousand streamlines per voxel were then initiated from each seed mask, with 0.5 step distance. A fiber was terminated when 1) it reached other seed masks; 2) it exceeded 2000 step limits; 3) it looped back to the same streamline; 4) its curvature exceeded 80 degree 5) it left the WM mask. Once the all fibers terminated, fibers that reached one of the seed masks were retained and counted to determine the connectivity between ROIs.

### 2.5. Individual-level Structural Brain Network Construction and Analyses

To construct the structural brain network for each subject, the 78 cortical and subcortical ROIs were used as network nodes. A pair of nodes were considered to have no anatomical connectivity (i.e., no edge in the network), if fiber tracts from neither of the two nodes successfully reached to the other one, during the probabilistic tractography step. The weight of a non-zero edge was first evaluated by averaging the number of fibers on both directions. This raw value was then transformed using logarithm function and normalized by dividing the maximum edge weight in the same network (Rubinov and Sporns, 2011). In addition, a non-zero edge was further set as zero if at least 60% of the whole study sample had a zero weight on this edge. This cutoff threshold was validated in previous studies for efficacy of controlling false positive and false negative rates of the generated connections (de Reus and van den Heuvel, 2013;Misic et al., 2018;Verhelst et al., 2018;Bathelt et al., 2019). Then for each subject, the 78 × 78 symmetric connectivity matrix was generated for construction of the weighted structural brain (**Figure 1F**).

The global and regional topological properties of the structural brain network from each subject were then estimated, including the network global and local efficiencies, network overall strength, and nodal global efficiency, nodal local efficiency, and nodal clustering coefficient of each node. All network topological property were calculated using Brain Connectivity Toolbox (Rubinov and Sporns, 2010).

The network global efficiency is a metric of the structural network integration that reflects the ability of information transferring across distributed brain areas (Latora and Marchiori, 2001). It was defined as

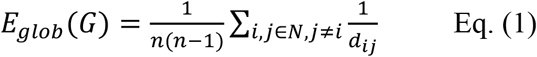

where *d*_*ij*_ was the inverse of the shortest distance between node *i* and *j* that was represented using the edge’s normalized weight. When two nodes were not directly connected, the shortest distance was the sum of the shortest connecting edges.

The network local efficiency estimates the network segregation and represents the fault tolerance level of the network (Latora and Marchiori, 2001), which was defined as

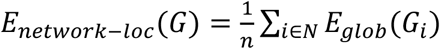

where *G*_*i*_ was the subnetwork consisted of all neighbor nodes of node *i*, and the global efficiency of subnetwork *G*_*i*_ is calculated using equation Eq. (1).

The network overall strength was defined as the average of the normalized weights of the edges in the network, which was used to represent the overall connectivity of the network.

The nodal global efficiency of node *i* is a measure of its nodal communication capacity with all other nodes in the network, which was defined as

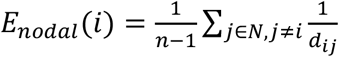

Nodal local efficiency of node *i* represents the robustness and integration of the subnetwork it belongs, which was defined as the global efficiency of the subnetwork consist of all the neighbors of *i*.

The nodal clustering coefficient describing the likelihood of whether the neighboring nodes of node *i* are interconnected with each other (Onnela et al., 2005). It was defined as

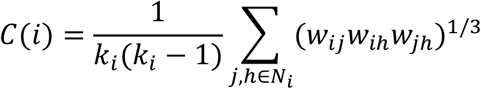

where *j, k* were neighbors of node *i*, and *k*_*i*_ was the number of neighbors of node *i*.

In a communicative network, there are certain nodes that have strong connections with other nodes, and/or frequently appear in the shortest between-node paths. These critical nodes are called “network hubs”, which serve to connect multiple segregated subnetworks and facilitate the intermodular integrations (Rubinov and Sporns, 2010). In our study, nodal strength and betweenness-centrality (BC) were estimated to characterize the hub property of each node in a network. The strength of a node was defined as the sum of the weights of its edges; whereas the BC attempted to measure the ability for one node to bridge indirectly connected nodes (Freeman, 1978). BC was defined as

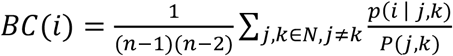

where *j, k* were node pairs in the network. *p*(*i* | *j, k*) was whether the shortest path between node *j* and node *k* passes through node *i. P*(*j, k*) was the total number of unique shortest path between node *j* and node *k*.

For each node in a WM structural brain network, its nodal global efficiency represent the integration of its associated WM structural subnetworks; whereas it nodal local efficiency and nodal clustering coefficient represent the modularity, and BC represents the connectivity of its associated WM subnetworks (Fagerholm et al., 2015;Jolly et al., 2020).

### 2.6. Group-level Analyses

Group statistics were carried out using SPSS 25 on macOS Mojave 10.14.1. Between-group comparisons in demographic, clinical, behavioral, and neurocognitive performance measures were conducted using chi-square test for categorical data (sex and ethics), and independent two sample t-test for numerical measures.

Group comparisons in the network topological measures were performed using a mixed-effects general linear model by setting TBI-A and controls as group variables, and adding IQ, age, SES as random-effect, and sex as fixed-effect covariates, respectively. In addition, the group-specific network hubs of each diagnostic group were examined using one-sample t test in the nodal strength and BC measures, respectively, with a threshold of 2 standard deviations higher than the group mean. Group comparisons in all of these network measures were controlled for potential multiple comparisons (in the total of 78 network nodes), using Bonferroni correction with a threshold of significance at corrected α ≤ 0.05 (Green and Diggle, 2007).

Brain-behavior associations in the TBI-A group were assessed using Pearson correlation between the T scores of the inattentive and hyperactive/impulsive subscales from Conners 3-PS and the network measures that showed significant between-group differences. The correlation analyses were controlled for potential multiple comparisons (in the total number of comparisons), by using Bonferroni correction with a threshold of significance at corrected α ≤ 0.05.

## 3. Results

### 3.1. Demographic, Clinical/Behavioral and Neurocognitive Performance Measures

As shown in **Table 1**, there were no significant between-group differences in the demographic and neurocognitive performance measures. Compared to the controls, the children with TBI-A showed significantly more inattentive (p<0.001) and hyperactive/impulsive (p<0.001) symptoms measured using the T scores in Conners 3-PS.

### 3.2. Topological Properties of the Structural Brain Network

The global network properties did not show significant between-group differences. Compared to controls, the TBI-A group showed significantly increased nodal local efficiency (*p*=0.0050) and nodal clustering coefficient (*p*=0.0009) in left inferior frontal gyrus; significantly increased BC (*p*=0.0374) in left superior frontal gyrus; and significantly increased nodal local efficiency (*p*=0.0361) and nodal clustering coefficient (*p*=0.0430) in right transverse temporal gyrus. Meanwhile, relative to controls, the TBI-A group also demonstrated significantly decreased nodal local efficiency (*p*=0.0264) in left parahippocampal gyrus; and greatly reduced nodal clustering coefficient (*p*=0.0173) in left supramarginal gyrus (**Table 2)**.

**Table 2.**
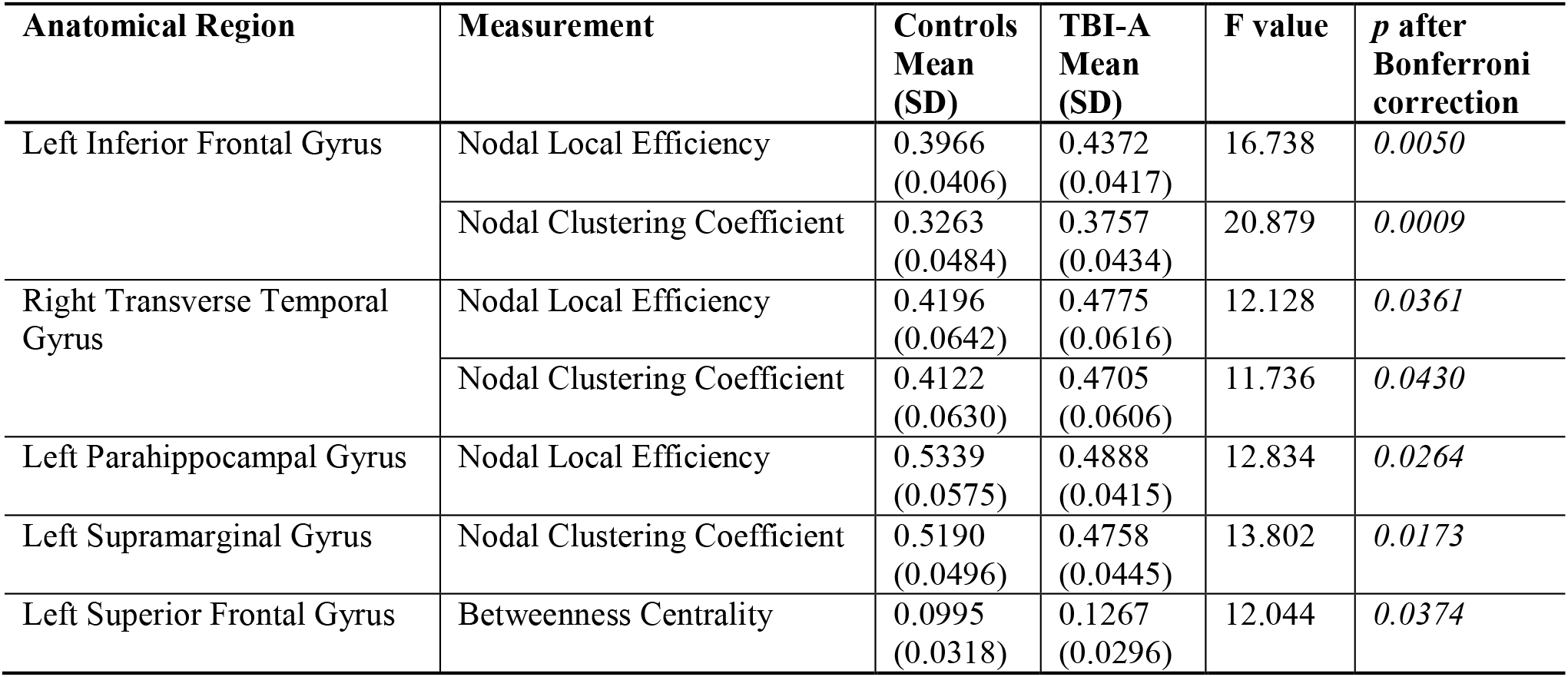
Anatomical regions that showed significant between-group differences in nodal topological properties of the structural brain network. TBI-A: TBI induced attention deficit; SD: Standard deviation.

In addition, distinct patterns of the within-group hub distribution were observed in the two diagnostic groups (**Figure 2**), with the precentral gyrus and putamen nucleus in the right hemisphere showing as hubs (measured by BC) in the TBI-A group but not in controls. Left precentral gyrus was identified as a hub in controls but not in the TBI-A group.

**Figure 2.**
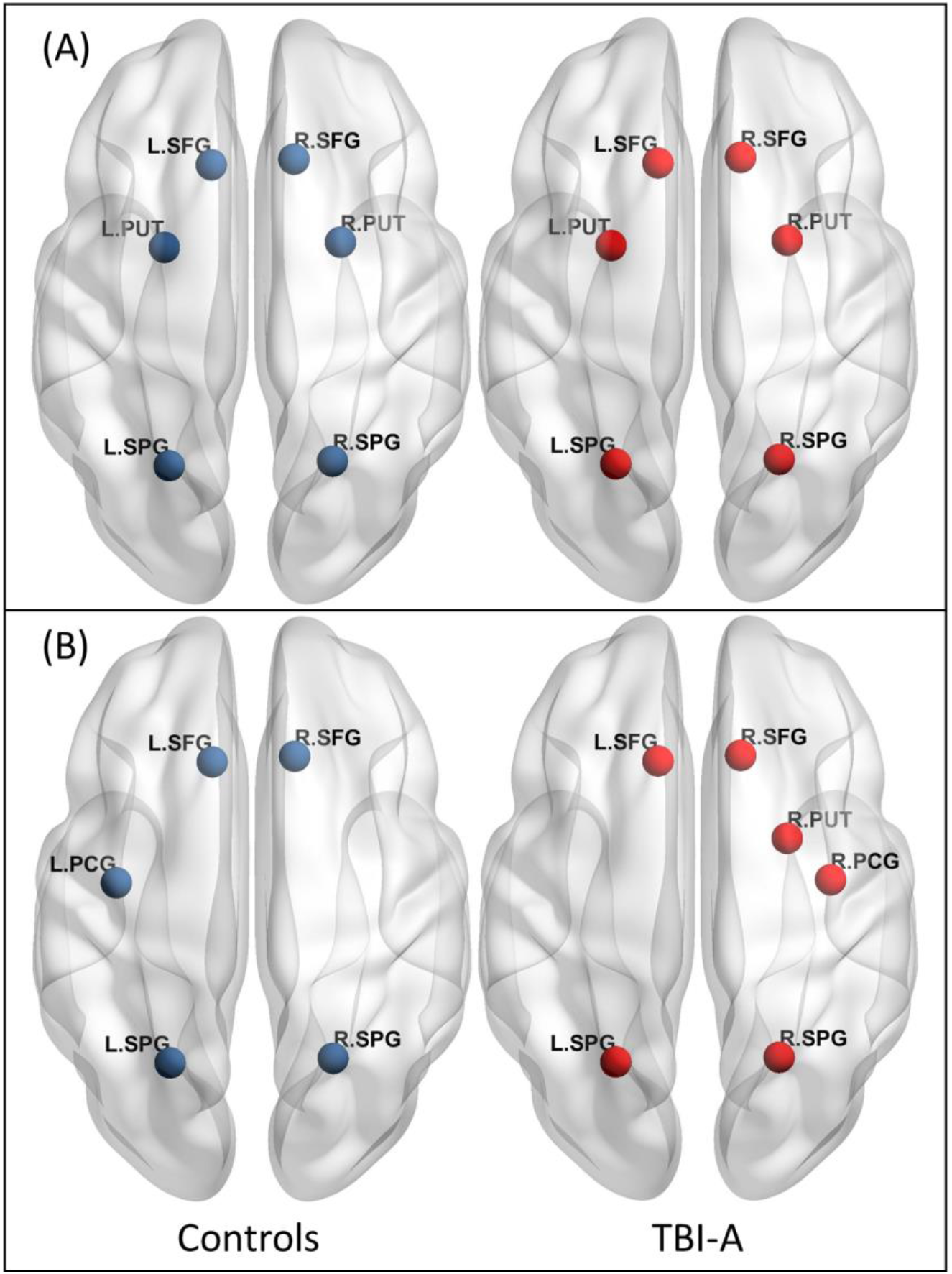
Network Hubs. (A) Nodal Strength defined network hubs, with controls on the left and TBI-A groups on the right. (A) Betweenness-centrality defined network hubs, with controls on the left and TBI-A groups on the right. Left precentral gyrus defined as a hub in controls. Right putamen and right precentral gyrus defined as hubs in TBI-A. TBI-A: TBI-induced attention deficits; SFG: Superior frontal gyrus; SPG: Superior parietal gyrus; PCG: Precentral gyrus; PUT: Putamen.

### 3.3. Brain-behavior Associations in the TBI-A Group

In the TBI-A group, increased nodal local efficiency of left parahippocampal gyrus was significantly associated with increased inattentive (r=0.405, *p*=0.024) and hyperactive/impulsive (r=0.457, *p*=0.01) symptoms; while greater nodal clustering coefficient of right transverse temporal gyrus was strongly associated with decreased hyperactive/impulsive symptoms (r=-0.468, *p*=0.008) (**Figure 3)**.

**Figure 3.**
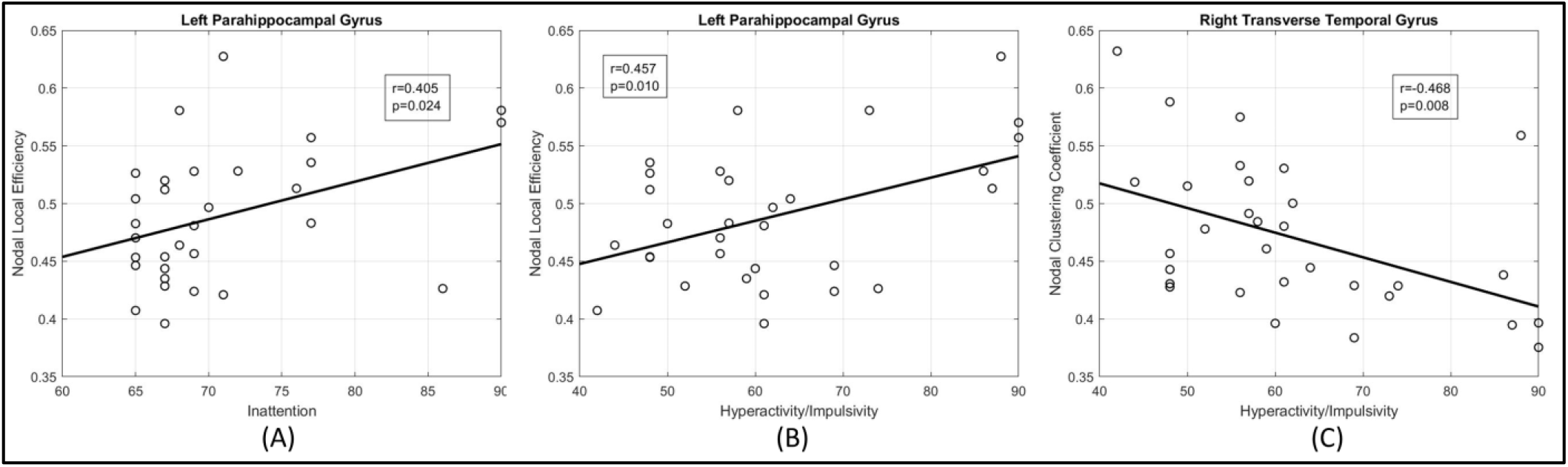
Brain-behavior Associations in the TBI-A Group. (A) Correlation of hyperactive/impulsive symptoms and nodal local efficiency of left parahippocampal gyrus. (B) Correlation of inattentive symptoms and nodal local efficiency of left parahippocampal gyrus. (C) Correlation of hyperactive/impulsive symptoms and nodal lustering coefficient of right transverse temporal gyrus.

## 4. Discussion

This study depicted for the first time that aberrant regional topological properties of the WM structural brain network play critical role in severe post-TBI attention deficits in children. Specifically, we found that relative to the group-matched controls, children with TBI-A had significantly increased nodal local efficiency and nodal clustering coefficient in the left inferior frontal gyrus, as well as significantly higher BC in the left superior frontal gyrus. These results suggest significantly increased structural connectivity and modularity of the subnetworks associated with left inferior and superior frontal gyri in children with severe post-TBI attention deficits. Chronic tissue abnormalities in children with mild and moderate TBI have been found to be mainly resulted from diffuse axonal injury (DAI), due to the abrupt stretching, twisting, and shearing of axons in the event of a mechanical blow (Roberts et al., 2016). Frontal lobe, located class to the anterior fossa of the skull, is one of the most vulnerable brain regions to DAI (Bigler, 2007). Existing neuroimaging studies in children with chronic TBI have consistently demonstrated structural anomalies in frontal cortex GM and the WM pathways connect it and other brain regions. For instance, multiple structural MRI and DTI studies have reported frontal GM volumetric reduction and cortical thinning (Wilde et al., 2012b;Bigler et al., 2013;Mayer et al., 2015;Dennis et al., 2016), as well as disrupted frontal WM integrity, represented by reduced WM FA and increased apparent diffusion coefficient in children with chronic TBI relative to group-matched controls (Wozniak et al., 2007;Wilde et al., 2011;Wilde et al., 2012a). Frontal tissue anomalies in children with TBI have also been found to link to long-term neurobehavioral impairments in domains such as executive control (Lipszyc et al., 2014) and learning and memory (Lindsey et al., 2019); whereas no evidence from previous quantitative clinical and neuroimaging studies have suggested strong associations between frontal GM/WM tissue alterations and post-TBI attention deficits in children. Along with these existing studies, results from the current study suggest that abnormal structural connectivity and modularity of the subnetworks associated with frontal lobe may be caused by TBI-induced structural damages in frontal cortex and associated WM structures; while these regional topological alterations of the WM structural network might not necessarily play the key role in long-term and severe post-TBI attention deficits in the affected individuals.

Compared to controls, the TBI-A group demonstrated significantly reduced nodal clustering coefficient in left supramarginal gyrus. This result of reduced topological modularity of the structural subnetwork in parietal regions is consistent with findings from a previous structural network study using deterministic tractography (Caeyenberghs et al., 2012). In addition, several previous DTI studies in children with chronic TBI have consistently reported significantly decreased FA of the superior longitudinal fasciculus, which is a major association tract that connects parietal lobe with frontal lobe (Ewing-Cobbs et al., 2016;Konigs et al., 2018;Molteni et al., 2019). An early structural MRI study reported significantly reduced cortical thickness of bilateral supramarginal gyri in children with chronic TBI children, relative to matched controls (Merkley et al., 2008); while a more recent longitudinal study reported significant associations between greater volume of left supramarginal gyrus and worse overall cognitive performance (Dennis et al., 2016). Meanwhile, task-based functional MRI studies have demonstrated abnormal supramarginal gyrus activation in children with chronic TBI, when performing a motor task (Caeyenberghs et al., 2009) and a working memory task (Newsome et al., 2008). However, similar to the fact from investigations in frontal regions, no evidence has yet suggested strong associations between parietal lobe GM/WM tissue alterations and post-TBI attention deficits in children.

Intriguingly, the current study found that relative to controls, the TBI-A group had significantly decreased nodal local efficiency in left parahippocampal gyrus and significantly increased nodal local efficiency and nodal clustering coefficient in right transverse temporal gyrus. Furthermore, nodal local efficiency in left parahippocampal gyrus showed significant positive correlations with the post-TBI inattentive and hyperactive symptoms, and nodal local efficiency in right transverse temporal gyrus showed significant negative correlations with the post-TBI hyperactive symptoms, in the group of TBI-A. Similar to the frontal lobe, temporal lobe is also among the most vulnerable brain regions for DAI, due to its anatomical location as the close proximity to the bony structure of the middle fossa of the skull (Bigler, 2007). TBI-related cortical GM atrophy and disrupted WM integrity in temporal lobe have been reported in several studies in children with chronic TBI (Wilde et al., 2005;Caeyenberghs et al., 2012;Wilde et al., 2012a;Dennis et al., 2016;Diez et al., 2017). The transverse temporal gyrus, also call Heschl’s gyrus, is the primary auditory cortex responsible for early processing related to speech understanding (Recanzone and Cohen, 2010;Arnott and Alain, 2011). It was also found to be part of the dorsal pathway in the bottom-up visual attention stream (Katsuki and Constantinidis, 2014), as well as subject to top-down influences of attention (Voisin et al., 2006). Parahippocampal gyrus belongs to the medial temporal system for visuospatial processing, which has intensive WM connections with frontal, parietal, occipital cortices, and midbrain structures. It was found to involve in selective attention during shifting and orienting processes through the ventral attention pathways (Corbetta and Shulman, 2002;Wager et al., 2004;Ochsner et al., 2012;Vossel et al., 2014). Both the transverse temporal and parahippocampal gyri are critical components in the multisensory integration system for attention processing (Cappe et al., 2009). These existing studies in cognitive neuroscience have provided strong scientific premise of our novel findings in the temporal lobe in children with TBI-A. Therefore, we suggest that TBI-related local re-modularity associated with the transverse temporal region, and structural segregation of the subnetworks connecting the parahippocampal gyrus with other brain regions, may have significant linkage with the onset of post-TBI inattentive and hyperactive/impulsive symptoms in children.

There are some limitations of the present study. First, the sample size is relatively small. Compared to other existing studies with similar sample sizes, the effect size of our study is larger, because of the inclusion criteria of the two diagnostic groups (the T-scores of inattentive and hyperactive subscales were ≥ 65 for TBI-A, while ≤60 for controls). The increased effect size can help improve statistical power of our study. Recently, several studies have reported effects of sex and SES on the long-term cognitive and behavioral outcomes in children with TBI (Yeates et al., 2012;Anderson et al., 2013;Scholten et al., 2015;Wade et al., 2016). Considering the relatively small sample size, we did not investigate the between-sex (male/female) topological differences of the structural brain network, and their interactions with the two diagnostic groups. Additional analyses of our sample did not show any trends of significant associations of the SES and time from injury with any clinical/behavioral measures in the TBI-A group. To partially remove the potential effects of these factors, we added sex as a fixed effect covariate, and SES as a random effect covariate, in the group-level analyses.

In summary, the current study demonstrated significantly altered regional topological organizations of the WM brain network in frontal, parietal, and temporal regions, in a more homogeneous subgroup of children with chronic TBI who had severe post-TBI attention deficits. The results further suggest that TBI-related WM structural re-modularity in the subnetworks associated with temporal lobe may significantly link to onset of severe post-TBI attention deficits in the affected children. These findings provide valuable implication for understanding the neurobiological substrates of TBI-A, and have the potential to serve as a biomarker guiding the development of more timely and tailored strategies for diagnoses and treatments to the affected individuals.

## Data Availability

Data are currently not available to public.

## Acknowledgements

Dr. Xiaobo Li designed the study. Meng Cao worked on literature searching, clinical and imaging data analyses, and wrote the first draft of the manuscript. Drs. Li, Haplerin, Mazzola, Catania, Goodman, Luo, Wu, and Alvarez edited and revised the manuscript. All authors contributed to and have approved the final manuscript.

## References

Adamson, C., Yuan, W., Babcock, L., Leach, J.L., Seal, M.L., Holland, S.K., and Wade, S.L. (2013). Diffusion tensor imaging detects white matter abnormalities and associated cognitive deficits in chronic adolescent TBI. Brain Inj 27, 454–463.

Anderson, V., Beauchamp, M.H., Yeates, K.O., Crossley, L., Hearps, S.J., and Catroppa, C. (2013). Social competence at 6 months following childhood traumatic brain injury. J Int Neuropsychol Soc 19, 539–550.

Andersson, J.L.R., and Sotiropoulos, S.N. (2016). An integrated approach to correction for off-resonance effects and subject movement in diffusion MR imaging. Neuroimage 125, 1063–1078.

Arnott, S.R., and Alain, C. (2011). The auditory dorsal pathway: orienting vision. Neurosci Biobehav Rev 35, 2162–2173.

Association, A.P. (2013). Diagnostic and statistical manual of mental disorders (DSM-5®). American Psychiatric Pub.

Backeljauw, B., and Kurowski, B.G. (2014). Interventions for attention problems after pediatric traumatic brain injury: what is the evidence? PM R 6, 814–824.

Bassett, D.S., Wymbs, N.F., Porter, M.A., Mucha, P.J., Carlson, J.M., and Grafton, S.T. (2011). Dynamic reconfiguration of human brain networks during learning. Proc Natl Acad Sci U S A 108, 7641–7646.

Bathelt, J., Johnson, A., Zhang, M., and Astle, D.E. (2019). The cingulum as a marker of individual differences in neurocognitive development. Sci Rep 9, 2281.

Baum, G.L., Ciric, R., Roalf, D.R., Betzel, R.F., Moore, T.M., Shinohara, R.T., Kahn, A.E., Vandekar, S.N.m Rupert, P.E., Quarmley, M., Cook, P.A., Elliott, M.A., Ruparel, K., Gur, R.E., Gur, R.C., Bassett, D.S., and Satterthwaite, T.D. (2017). Modular Segregation of Structural Brain Networks Supports the Development of Executive Function in Youth. Curr Biol 27, 1561–1572 e1568.

Behrens, T.E., Berg, H.J., Jbabdi, S., Rushworth, M.F., and Woolrich, M.W. (2007). Probabilistic diffusion tractography with multiple fibre orientations: What can we gain? Neuroimage 34, 144–155.

Bigler, E.D. (2007). Anterior and middle cranial fossa in traumatic brain injury: relevant neuroanatomy and neuropathology in the study of neuropsychological outcome. Neuropsychology 21, 515–531.

Bigler, E.D., Abildskov, T.J., Petrie, J., Farrer, T.J., Dennis, M., Simic, N., Taylor, H.G., Rubin, K.H., Vannatta, K., Gerhardt, C.A., Stancin, T., and Owen Yeates, K. (2013). Heterogeneity of brain lesions in pediatric traumatic brain injury. Neuropsychology 27, 438–451.

Blakemore, S.J., and Choudhury, S. (2006). Development of the adolescent brain: implications for executive function and social cognition. J Child Psychol Psychiatry 47, 296–312.

Caeyenberghs, K., Leemans, A., De Decker, C., Heitger, M., Drijkoningen, D., Linden, C.V., Sunaert, S., and Swinnen, S.P. (2012). Brain connectivity and postural control in young traumatic brain injury patients: A diffusion MRI based network analysis. Neuroimage Clin 1, 106–115.

Caeyenberghs, K., Wenderoth, N., Smits-Engelsman, B.C., Sunaert, S., and Swinnen, S.P. (2009). Neural correlates of motor dysfunction in children with traumatic brain injury: exploration of compensatory recruitment patterns. Brain 132, 684–694.

Cappe, C., Rouiller, E.M., and Barone, P. (2009). Multisensory anatomical pathways. Hear Res 258, 28–36.

Carskadon, M.A., and Acebo, C. (1993). A self-administered rating scale for pubertal development. J Adolesc Health 14, 190–195.

Chu, S.H., Parhi, K.K., and Lenglet, C. (2018). Function-specific and Enhanced Brain Structural Connectivity Mapping via Joint Modeling of Diffusion and Functional MRI. Sci Rep 8, 4741. Conners, C.K. (Year). “Conners 3”: MHS).

Corbetta, M., and Shulman, G.L. (2002). Control of goal-directed and stimulus-driven attention in the brain. Nat Rev Neurosci 3, 201–215.

De Reus, M.A., and Van Den Heuvel, M.P. (2013). Estimating false positives and negatives in brain networks. Neuroimage 70, 402–409.

Dennis, E.L., Hua, X., Villalon-Reina, J., Moran, L.M., Kernan, C., Babikian, T., Mink, R., Babbitt, C., Johnson, J., Giza, C.C., Thompson, P.M., and Asarnow, R.F. (2016). Tensor-Based Morphometry Reveals Volumetric Deficits in Moderate=Severe Pediatric Traumatic Brain Injury. J Neurotrauma 33, 840–852.

Dennis, E.L., Jin, Y., Villalon-Reina, J.E., Zhan, L., Kernan, C.L., Babikian, T., Mink, R.B., Babbitt, C.J., Johnson, J.L., Giza, C.C., Thompson, P.M., and Asarnow, R.F. (2015). White matter disruption in moderate/severe pediatric traumatic brain injury: advanced tract-based analyses. Neuroimage Clin 7, 493–505.

Dewan, M.C., Mummareddy, N., Wellons, J.C., 3rd, and Bonfield, C.M. (2016). Epidemiology of Global Pediatric Traumatic Brain Injury: Qualitative Review. World Neurosurg 91, 497–509 e491.

Diez, I., Drijkoningen, D., Stramaglia, S., Bonifazi, P., Marinazzo, D., Gooijers, J., Swinnen, S.P., and Cortes, J.M. (2017). Enhanced prefrontal functional-structural networks to support postural control deficits after traumatic brain injury in a pediatric population. Netw Neurosci 1, 116–142.

Eckner, J.T., Kutcher, J.S., and Richardson, J.K. (2011). Effect of concussion on clinically measured reaction time in 9 NCAA division I collegiate athletes: a preliminary study. PM R 3, 212–218.

Ewing-Cobbs, L., Johnson, C.P., Juranek, J., Demaster, D., Prasad, M., Duque, G., Kramer, L., Cox, C.S., and Swank, P.R. (2016). Longitudinal diffusion tensor imaging after pediatric traumatic brain injury: Impact of age at injury and time since injury on pathway integrity. Hum Brain Mapp 37, 3929–3945.

Ewing-Cobbs, L., Prasad, M.R., Swank, P., Kramer, L., Cox, C.S., Jr., Fletcher, J.M., Barnes, M., Zhang, X., and Hasan, K.M. (2008). Arrested development and disrupted callosal microstructure following pediatric traumatic brain injury: relation to neurobehavioral outcomes. Neuroimage 42, 1305–1315.

Faber, J., Wilde, E.A., Hanten, G., Ewing-Cobbs, L., Aitken, M.E., Yallampalli, R., Macleod, M.C., Mullins, S.H., Chu, Z.D., Li, X., Hunter, J.V., Noble-Haeusslein, L., and Levin, H.S. (2016). Ten-year outcome of early childhood traumatic brain injury: Diffusion tensor imaging of the ventral striatum in relation to executive functioning. Brain Inj 30, 1635–1641.

Fagerholm, E.D., Hellyer, P.J., Scott, G., Leech, R., and Sharp, D.J. (2015). Disconnection of network hubs and cognitive impairment after traumatic brain injury. Brain 138, 1696–1709.

Fischl, B. (2012). FreeSurfer. Neuroimage 62, 774–781.

Freeman, L.C.J.S.N. (1978). Centrality in social networks conceptual clarification. 1, 215–239.

Green, G.H., and Diggle, P.J. (2007). On the operational characteristics of the Benjamini and Hochberg false discovery rate procedure. Statistical applications in genetics and molecular biology 6.

Imms, P., Clemente, A., Cook, M., D’souza, W., Wilson, P.H., Jones, D.K., and Caeyenberghs, K. (2019). The structural connectome in traumatic brain injury: A meta-analysis of graph metrics. Neurosci Biobehav Rev 99, 128–137.

Jancke, L., Shah, N.J., Posse, S., Grosse-Ryuken, M., and Muller-Gartner, H.W. (1998). Intensity coding of auditory stimuli: an fMRI study. Neuropsychologia 36, 875–883.

Jenkinson, M., Beckmann, C.F., Behrens, T.E., Woolrich, M.W., and Smith, S.M. (2012). Fsl. Neuroimage 62, 782–790.

Jolly, A.E., Scott, G.T., Sharp, D.J., and Hampshire, A.H. (2020). Distinct patterns of structural damage underlie working memory and reasoning deficits after traumatic brain injury. Brain 143, 1158–1176.

Katsuki, F., and Constantinidis, C. (2014). Bottom-up and top-down attention: different processes and overlapping neural systems. Neuroscientist 20, 509–521.

Kaufman, J., Birmaher, B., Brent, D.A., Ryan, N.D., and Rao, U. (2000). K-Sads-Pl.

Konigs, M., Heij, H.A., Van Der Sluijs, J.A., Vermeulen, R.J., Goslings, J.C., Luitse, J.S., Poll-The, B.T., Beelen, A., Van Der Wees, M., Kemps, R.J., Catsman-Berrevoets, C.E., and Oosterlaan, J. (2015). Pediatric Traumatic Brain Injury and Attention Deficit. Pediatrics 136, 534–541.

Konigs, M., Pouwels, P.J., Ernest Van Heurn, L.W., Bakx, R., Jeroen Vermeulen, R., Goslings, J.C., Carel Goslings, J., Poll-The, B.T., Van Der Wees, M., Catsman-Berrevoets, C.E., and Oosterlaan, J. (2018). Relevance of neuroimaging for neurocognitive and behavioral outcome after pediatric traumatic brain injury. Brain Imaging Behav 12, 29–43.

Konigs, M., Van Heurn, L.W.E., Bakx, R., Vermeulen, R.J., Goslings, J.C., Poll-The, B.T., Van Der Wees, M., Catsman-Berrevoets, C.E., Oosterlaan, J., and Pouwels, P.J.W. (2017). The structural connectome of children with traumatic brain injury. Hum Brain Mapp 38, 3603–3614.

Kramer, M.E., Chiu, C.Y., Walz, N.C., Holland, S.K., Yuan, W., Karunanayaka, P., and Wade, S.L. (2008). Long-term neural processing of attention following early childhood traumatic brain injury: fMRI and neurobehavioral outcomes. J Int Neuropsychol Soc 14, 424–435.

Kurowski, B., Wade, S.L., Cecil, K.M., Walz, N.C., Yuan, W., Rajagopal, A., and Holland, S.K. (2009). Correlation of diffusion tensor imaging with executive function measures after early childhood traumatic brain injury. J Pediatr Rehabil Med 2, 273–283.

Kurowski, B.G., Epstein, J.N., Pruitt, D.W., Horn, P.S., Altaye, M., and Wade, S.L. (2019). Benefits of Methylphenidate for Long-Term Attention Problems After Traumatic Brain Injury in Childhood: A Randomized, Double-Masked, Placebo-Controlled, Dose-Titration, Crossover Trial. J Head Trauma Rehabil 34, E1–E12.

Latora, V., and Marchiori, M. (2001). Efficient behavior of small-world networks. Phys Rev Lett 87, 198701.

Le Fur, C., Camara-Costa, H., Francillette, L., Opatowski, M., Toure, H., Brugel, D., Laurent-Vannier, A., Meyer, P., Watier, L., Dellatolas, G., and Chevignard, M. (2019). Executive functions and attention 7years after severe childhood traumatic brain injury: Results of the Traumatisme Grave de l’Enfant (TGE) cohort. Ann Phys Rehabil Med.

Leblond, E., Smith-Paine, J., Riemersma, J.J., Horn, P.S., Wade, S.L., and Kurowski, B.G. (2019). Influence of Methylphenidate on Long-Term Neuropsychological and Everyday Executive Functioning After Traumatic Brain Injury in Children with Secondary Attention Problems. J Int Neuropsychol Soc 25, 740–749.

Lindsey, H.M., Lalani, S.J., Mietchen, J., Gale, S.D., Wilde, E.A., Faber, J., Macleod, M.C., Hunter, J.V., Chu, Z.D., Aitken, M.E., Ewing-Cobbs, L., and Levin, H.S. (2019). Acute pediatric traumatic brain injury severity predicts long-term verbal memory performance through suppression by white matter integrity on diffusion tensor imaging. Brain Imaging Behav.

Lipszyc, J., Levin, H., Hanten, G., Hunter, J., Dennis, M., and Schachar, R. (2014). Frontal white matter damage impairs response inhibition in children following traumatic brain injury. Arch Clin Neuropsychol 29, 289–299.

Lumba-Brown, A., Yeates, K.O., Sarmiento, K., Breiding, M.J., Haegerich, T.M., Gioia, G.A., Turner, M., Benzel, E.C., Suskauer, S.J., Giza, C.C., Joseph, M., Broomand, C., Weissman, B., Gordon, W., Wright, D.W., Moser, R.S., Mcavoy, K., Ewing-Cobbs, L., Duhaime, A.C., Putukian, M.,Holshouser, B., Paulk, D., Wade, S.L., Herring, S.A., Halstead, M., Keenan, H.T., Choe, M., Christian, C.W., Guskiewicz, K., Raksin, P.B., Gregory, A., Mucha, A., Taylor, H.G., Callahan, J.M., Dewitt, J., Collins, M.W., Kirkwood, M.W., Ragheb, J., Ellenbogen, R.G., Spinks, T.J., Ganiats, T.G., Sabelhaus, L.J., Altenhofen, K., Hoffman, R., Getchius, T., Gronseth, G., Donnell, Z., O’connor, R.E., and Timmons, S.D. (2018). Diagnosis and Management of Mild Traumatic Brain Injury in Children: A Systematic Review. JAMA Pediatr 172, e182847.

Max, J.E., Schachar, R.J., Levin, H.S., Ewing-Cobbs, L., Chapman, S.B., Dennis, M., Saunders, A., Landis, J.J.J.O.T.a.a.O.C., and Psychiatry, A. (2005). Predictors of secondary attentiondeficit/hyperactivity disorder in children and adolescents 6 to 24 months after traumatic brain injury. 44, 1041–1049.

Mayer, A.R., Hanlon, F.M., and Ling, J.M. (2015). Gray matter abnormalities in pediatric mild traumatic brain injury. J Neurotrauma 32, 723–730.

Merkley, T.L., Bigler, E.D., Wilde, E.A., Mccauley, S.R., Hunter, J.V., and Levin, H.S. (2008). Diffuse changes in cortical thickness in pediatric moderate-to-severe traumatic brain injury. J Neurotrauma 25, 1343–1345.

Misic, B., Betzel, R.F., Griffa, A., De Reus, M.A., He, Y., Zuo, X.N., Van Den Heuvel, M.P., Hagmann, P., Sporns, O., and Zatorre, R.J. (2018). Network-Based Asymmetry of the Human Auditory System. Cereb Cortex 28, 2655–2664.

Molteni, E., Pagani, E., Strazzer, S., Arrigoni, F., Beretta, E., Boffa, G., Galbiati, S., Filippi, M., and Rocca, M.A. (2019). Fronto-temporal vulnerability to disconnection in paediatric moderate and severe traumatic brain injury. Eur J Neurol 26, 1183–1190.

Narad, M.E., Riemersma, J., Wade, S.L., Smith-Paine, J., Morrison, P., Taylor, H.G., Yeates, K.O., and Kurowski, B.G. (2019). Impact of Secondary ADHD on Long-Term Outcomes After Early Childhood Traumatic Brain Injury. J Head Trauma Rehabil.

Newsome, M.R., Steinberg, J.L., Scheibel, R.S., Troyanskaya, M., Chu, Z., Hanten, G., Lu, H., Lane, S., Lin, X., Hunter, J.V., Vasquez, C., Zientz, J., Li, X., Wilde, E.A., and Levin, H.S. (2008). Effects of traumatic brain injury on working memory-related brain activation in adolescents. Neuropsychology 22, 419–425.

Ochsner, K.N., Silvers, J.A., and Buhle, J.T. (2012). Functional imaging studies of emotion regulation: a synthetic review and evolving model of the cognitive control of emotion. Ann N Y Acad Sci 1251, E1–24.

Oldfield, R.C. (1971). The assessment and analysis of handedness: the Edinburgh inventory. Neuropsychologia 9, 97–113.

Onnela, J.P., Saramaki, J., Kertesz, J., and Kaski, K. (2005). Intensity and coherence of motifs in weighted complex networks. Phys Rev E Stat Nonlin Soft Matter Phys 71, 065103.

Polinder, S., Haagsma, J.A., Van Klaveren, D., Steyerberg, E.W., and Van Beeck, E.F. (2015). Healthrelated quality of life after TBI: a systematic review of study design, instruments, measurement properties, and outcome. Popul Health Metr 13, 4.

Recanzone, G.H., and Cohen, Y.E. (2010). Serial and parallel processing in the primate auditory cortex revisited. Behav Brain Res 206, 1-7.

Roberts, R.M., Mathias, J.L., and Rose, S.E. (2016). Relationship Between Diffusion Tensor Imaging (DTI) Findings and Cognition Following Pediatric TBI: A Meta-Analytic Review. Dev Neuropsychol 41, 176–200.

Rubinov, M., and Sporns, O. (2010). Complex network measures of brain connectivity: uses and interpretations. Neuroimage 52, 1059–1069.

Rubinov, M., and Sporns, O. (2011). Weight-conserving characterization of complex functional brain networks. Neuroimage 56, 2068–2079.

Ryan, N.P., Genc, S., Beauchamp, M.H., Yeates, K.O., Hearps, S., Catroppa, C., Anderson, V.A., and Silk, T.J. (2018). White matter microstructure predicts longitudinal social cognitive outcomes after paediatric traumatic brain injury: a diffusion tensor imaging study. Psychol Med 48, 679–691.

Scholten, A.C., Haagsma, J.A., Andriessen, T.M., Vos, P.E., Steyerberg, E.W., Van Beeck, E.F., and Polinder, S. (2015). Health-related quality of life after mild, moderate and severe traumatic brain injury: patterns and predictors of suboptimal functioning during the first year after injury. Injury 46, 616–624.

Spreng, R.N., Sepulcre, J., Turner, G.R., Stevens, W.D., and Schacter, D.L. (2013). Intrinsic architecture underlying the relations among the default, dorsal attention, and frontoparietal control networks of the human brain. J Cogn Neurosci 25, 74–86.

Strazzer, S., Rocca, M.A., Molteni, E., De Meo, E., Recla, M., Valsasina, P., Arrigoni, F., Galbiati, S., Bardoni, A., and Filippi, M. (2015). Altered Recruitment of the Attention Network Is Associated with Disability and Cognitive Impairment in Pediatric Patients with Acquired Brain Injury. Neural Plast 2015, 104282.

Teasdale, G., and Jennett, B. (1974). Assessment of coma and impaired consciousness. A practical scale. Lancet 2, 81–84.

Tlustos, S.J., Chiu, C.Y., Walz, N.C., Holland, S.K., Bernard, L., and Wade, S.L. (2011). Neural correlates of interference control in adolescents with traumatic brain injury: functional magnetic resonance imaging study of the counting stroop task. J Int Neuropsychol Soc 17, 181–189.

Treble, A., Hasan, K.M., Iftikhar, A., Stuebing, K.K., Kramer, L.A., Cox, C.S., Jr., Swank, P.R., and Ewing-Cobbs, L. (2013). Working memory and corpus callosum microstructural integrity after pediatric traumatic brain injury: a diffusion tensor tractography study. J Neurotrauma 30, 1609–1619.

Verhelst, H., Vander Linden, C., De Pauw, T., Vingerhoets, G., and Caeyenberghs, K. (2018). Impaired rich club and increased local connectivity in children with traumatic brain injury: Local support for the rich? Hum Brain Mapp 39, 2800–2811.

Voisin, J., Bidet-Caulet, A., Bertrand, O., and Fonlupt, P. (2006). Listening in silence activates auditory areas: a functional magnetic resonance imaging study. J Neurosci 26, 273–278.

Vossel, S., Geng, J.J., and Fink, G.R. (2014). Dorsal and ventral attention systems: distinct neural circuits but collaborative roles. Neuroscientist 20, 150–159.

Wade, S.L., Zhang, N., Yeates, K.O., Stancin, T., and Taylor, H.G. (2016). Social Environmental Moderators of Long-term Functional Outcomes of Early Childhood Brain Injury. JAMA Pediatr 170, 343–349.

Wager, T.D., Jonides, J., and Reading, S. (2004). Neuroimaging studies of shifting attention: a metaanalysis. Neuroimage 22, 1679–1693.

Watson, C.G., Demaster, D., and Ewing-Cobbs, L. (2019). Graph theory analysis of DTI tractography in children with traumatic injury. Neuroimage Clin 21, 101673.

Wechsler, D. (2011). Wechsler Abbreviated Scale of Intelligence–Second Edition (WASI-II). NCS Pearson.

Wilde, E.A., Ayoub, K.W., Bigler, E.D., Chu, Z.D., Hunter, J.V., Wu, T.C., Mccauley, S.R., and Levin, H.S. (2012a). Diffusion tensor imaging in moderate-to-severe pediatric traumatic brain injury: changes within an 18 month post-injury interval. Brain Imaging Behav 6, 404–416.

Wilde, E.A., Hunter, J.V., Newsome, M.R., Scheibel, R.S., Bigler, E.D., Johnson, J.L., Fearing, M.A., Cleavinger, H.B., Li, X., Swank, P.R., Pedroza, C., Roberson, G.S., Bachevalier, J., and Levin, H.S.(2005). Frontal and temporal morphometric findings on MRI in children after moderate to severe traumatic brain injury. J Neurotrauma 22, 333–344.

Wilde, E.A., Merkley, T.L., Bigler, E.D., Max, J.E., Schmidt, A.T., Ayoub, K.W., Mccauley, S.R., Hunter, J.V., Hanten, G., Li, X., Chu, Z.D., and Levin, H.S. (2012b). Longitudinal changes in cortical thickness in children after traumatic brain injury and their relation to behavioral regulation and emotional control. Int J Dev Neurosci 30, 267–276.

Wilde, E.A., Newsome, M.R., Bigler, E.D., Pertab, J., Merkley, T.L., Hanten, G., Scheibel, R.S., Li, X., Chu, Z., Yallampalli, R., Hunter, J.V., and Levin, H.S. (2011). Brain imaging correlates of verbal working memory in children following traumatic brain injury. Int J Psychophysiol 82, 86–96.

Wozniak, J.R., Krach, L., Ward, E., Mueller, B.A., Muetzel, R., Schnoebelen, S., Kiragu, A., and Lim, K.O. (2007). Neurocognitive and neuroimaging correlates of pediatric traumatic brain injury: a diffusion tensor imaging (DTI) study. Arch Clin Neuropsychol 22, 555–568.

Yeates, K.O., Taylor, H.G., Rusin, J., Bangert, B., Dietrich, A., Nuss, K., and Wright, M. (2012). Premorbid child and family functioning as predictors of post-concussive symptoms in children with mild traumatic brain injuries. Int J Dev Neurosci 30, 231–237.

Yuan, W., Treble-Barna, A., Sohlberg, M.M., Harn, B., and Wade, S.L. (2017). Changes in Structural Connectivity Following a Cognitive Intervention in Children With Traumatic Brain Injury. Neurorehabil Neural Repair 31, 190–201.

